# Correcting the Reproduction Number for Time-Varying Tests: a Proposal and an Application to COVID-19 in France*

**DOI:** 10.1101/2020.12.01.20241570

**Authors:** Christelle Baunez, Mickaël Degoulet, Stéphane Luchini, Matteo L. Pintus, Patrick A. Pintus, Miriam Teschl

## Abstract

We provide a novel way to correct the effective reproduction number for the time-varying amount of tests, using the acceleration index (Baunez et al., 2021) as a simple measure of viral spread dynamics. Not correcting results in the reproduction number being a biased estimate of viral acceleration and we provide a formal decomposition of the resulting bias, involving the useful notions of test and infectivity intensities. When applied to French data for the COVID-19 pandemic (May 13, 2020 - October 26, 2022), our decomposition shows that the reproduction number, when considered alone, characteristically underestimates the resurgence of the pandemic, compared to the acceleration index which accounts for the time-varying volume of tests. Because the acceleration index aggregates all relevant information and captures in real time the sizable time variation featured by viral circulation, it is a more parsimonious indicator to track the dynamics of an infectious disease outbreak in real time, compared to the equivalent alternative which would combine the reproduction number with the test and infectivity intensities.

**JEL Classification Numbers:** I18; H12

## 1 Introduction

The reproduction number is a widely used measure of how fast a pathogen propagates both at the outset and during an infectious disease outbreak (see for example May and Anderson [20]). One of its major shortcomings, however, is that it does not control for the quantity of tests (or any diagnostics) performed in real time. Doing so is of crucial importance for two reasons. One is the fact that accurate empirical estimates of reproduction numbers are time-varying in nature (see e.g. Fraser [13] among many others). A considerable source of time variation comes from the fact that the amount of tests changes substantially across time and hence affects the number of known cases, due to demand and supply effects. Second, testing acts as a magnifying lens on viral activity at least on the part of the population that has effectively been tested. The reproduction number however does not rely on that information but rather makes assumptions on infectivity, based on the observation of onset of symptoms and transmission in closed systems such as households (see e.g. Cori et al. [10]). But this information, again, depends on tests (or any diagnostics more generally). Hence inferring infectivity from (assumed) transmissions is only secondary information based on the availability of tests.

Soon after the onset of COVID-19, it was possible to diagnose people using PCR tests. From a public health perspective, this is of course a much more favorable situation, compared to other infectious diseases for which biological tests are either nonexistent or available much later after the disease has been discovered. Widespread testing allows early care and treatment whenever diagnostics are performed. In addition, it is a rather trivial observation that whenever information about how many tests are performed in a given period is available, that information should be used to assess the dynamics of viral spread. After all, positive and negative cases do not fall from heaven; quite to the contrary, they become visible only through the lens of testing. Considering only positive cases but ignoring tests means ignoring also how many negative cases are out there, which is the flip side of the disease and informs about how many people are not infected in a given population. The latter is a relevant and useful piece of information since incidence of the infectious disease should ideally be measured against the tested population, not the entire population which includes people with unknown health status. Given that it is hardly possible to think of reasons that would justify ignoring deliberately such data about the extent of testing, the question then becomes how to incorporate that bit of evidence into any indicator that aims at tracking the dynamics of viral spread. This is the core question that we address in this paper.

In Baunez et al. [4, 5, 6], we have introduced an alternative and novel measure of viral spread in the context of COVID-19 - the acceleration index. This measure considers the variation of cases *relative to* the variation of tests and thus avoids the shortcomings and addresses the important question mentioned above. The purpose of this article is to discuss the reproduction number in the light of our acceleration index, and to show that the former is actually a special case of the latter and in fact a less accurate metric of the pandemic’s time-varying spread.

We examine this important issue in two steps. In Section 2, we start from the very definition of the reproduction number as a gross rate of growth of infected people, traditionally denoted *R*, and derive a general formula that connects it to our acceleration index that we denote *ε*. The acceleration index is an elasticity that measures the proportional responsiveness of cases to tests, and it can also be thought of as the ratio between the current and average viral speeds. More specifically, we present an explicit measure of the ratio between *R* and *ε*, the interpretation of which is further discussed in terms of the infectivity and test intensities. Our theoretical inquiry stresses that while the acceleration index is a ratio of growth rates - that of cases divided by that of tests - the reproduction number tracks only the growth rate of cases. In other words, the acceleration index corrects the reproduction number for the time-varying amount of tests. Not doing so results in the reproduction number being a biased estimate of viral acceleration and we provide a formal decomposition of the resulting bias. The main conclusion we derive is that the reproduction number tends to overestimate (respectively underestimate) the dynamics of viral spread compared to the acceleration index when the amount of tests is large enough (respectively small enough).

In Section 3, we apply such an analysis to France, using an exhaustive data-set covering May 13, 2020, to October 26, 2022, and including pre- and post-vaccination periods. We show that there is a sizeable difference between both measures. Indeed, the reproduction number *R* largely *underestimates* the spread of the virus, compared to our test-controlled measure of viral acceleration. This discrepancy is particularly severe for the epidemic wave due to the Omicron strains during automn 2022. In Appendix C, we provide a similar analysis for five other countries to show that this result is not limited to the French case. It is in this sense that we say that the reproduction number is biased when tests are time-varying. This has obviously important consequences if the reproduction number is used as the basis for public health decisions such as entering or exiting a lock-down. We also look at the effects of the second lock-down period in France, which started October 30, 2020, through the lens of both indicators, as a further example that illustrates the bias unavoidably implied by not adjusting for the volume of tests over time when measuring the pandemic’s acceleration.

A key conclusion follows from our theoretical and empirical results. If public health authorities aim at measuring as accurately as possible viral acceleration, they have to rely on one of the following strategies: track in real time either the acceleration index alone, or a combination of the reproduction number together with the test and infectivity intensities. Although both strategies are formally equivalent, the latter is not only less parsimonious, it is also arguably more delicate to operate in practice since one would then like to control the bias that inevitably comes from time-varying tests. This is one of the main reasons why we argue in favor of using the acceleration index.

## 2 Materials and Methods

About a century ago, a series of seminal articles by Kermack and McKendrick [15, 16, 17] have laid the foundations for a mathematical theory of epidemics. More specifically, their compartmental (that is, ***S****usceptible*, ***I****nfected* and ***R****emoved* or SIR-type) and time-since-infection models have been extensively used and refined in the academic literature about infectious and emerging diseases. A core concept in this paradigm is the reproduction number, usually noted *R*, which roughly captures how many secondary cases originate, on average, from a pool of primary cases who is still currently infectious (see May and Anderson [20]).

As evident from publications by health agencies around the world since the onset of COVID-19, much of the guidance for designing policy measures to curb the pandemic relies prominently on estimates of *R*, among other things. The reproduction number is initially a theoretical concept, conceived to understand the transmissibility of an epidemic. Many efforts have been put into defining ways to empirically estimate it. Broadly speaking, estimation strategies fall into two broad categories. The first one rests on the basic SIR model (see e.g. Weiss [25] for a clear exposition), which predicts that the reproduction number *R* is the product of four parameters: the duration of infection, the number of contacts per case and the fraction of contacts who are in turn infected, on average, and finally the fraction of total population susceptible to infection. Although each of these parameters could be estimated in real-time, this turns out to be a gigantic task, in particular when a novel pathogen like SARS-Cov-2 emerges. A short-cut to avoid such a demanding procedure is to fit a SIR model using the number of cases, so as to estimate *R* directly, given the infection duration (see, among many others, Althaus [1] for a recent example related to Ebola using maximum likelihood estimation). This is feasible, even in real-time, provided that enough data is available to ensure precision and structural assumptions about the time-dependency of *R* are made. A caveat, though, is that such fitting procedures have limitations (see e.g. Cori et al. [11]). An additional issue arising from estimation based on compartmental models is the sizeable range of estimates. See Chris et al. [2] for SARS, and Viceconte and Petrosillo [23] for the early stages of COVID-19. The second estimation strategy addresses more directly the time-varying dimension of *R*, which is more in line with epidemiological and clinical data. Many health agencies rely on such estimates of time-since-infection transmission models rather than SIR-type models. Here the basic idea is that *R* is essentially (1+) the growth rate of infected, which is the ratio between the number of new (that is, secondary) cases arising, say, within 24 hours, and the number of primary cases (see Fraser [13]). For example, the French agency in charge of health statistics uses the Cori method, after Cori et al. [10] (see https://www.santepubliquefrance.fr/content/download/266456/2671953). Other European health agencies are also using this method, e.g. Austria (see https://www.ages.at/download/0/0/e03842347d92e5922e76993df9ac8e9b28635caa/fileadmin/AGES2015/Wissen-Aktuell/COVID19/Methoden_zur_Schätzung_der_epi_Parameter.pdf) and Germany (see https://www.rki.de/DE/Content/InfAZ/N/Neuartiges_Coronavirus/Projekte_RKI/R-Wert-Erlaeuterung.pdf?__blob=publicationFile).

Our task here is to relate the acceleration index defined in Baunez et al. [5, 4] and the reproduction number that is estimated using the time-since-infection approach just described. The main purpose of this section is to derive a theoretical relationship between both concepts, which helps both to explain why they are different, to give a sense of the magnitude of their difference, and to state the conditions under which they are equivalent. We then turn, in the next section, to data to gauge whether the difference between the two matters to track the COVID-19 pandemic.

Suppose that data is available about the number of tested and positive persons, up to end date *T*. Denote {*p*_1_, …, *p*_*T*_} the historical times series of the new (per period) number of positive persons from date *t* = 1 to end date *t* = *T*. Similarly, {*d*_1_, …, *d*_*T*_} is the historical times series of new (per period) diagnosed/tested persons. Denote 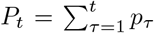 and 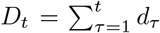 the cumulative numbers of positive and diagnosed persons up to date *t*.

As stressed in Baunez et al. [5], accurate information about the dynamics of a pandemic rests on both the number of cases and the number of tests, and the former cannot be properly understood without the latter. In that paper, we introduce an acceleration index, denoted *ε*_*T*_ at date *T*, which is an elasticity that measures the proportional responsiveness of cases with respect to tests. Given that the number of cases and tests are not necessarily varying at the same rate across time, groups and also space, the acceleration index measures the percentage change of cases with respect to a percentage change of tests and is thus unit-free. The acceleration index is defined as follows:

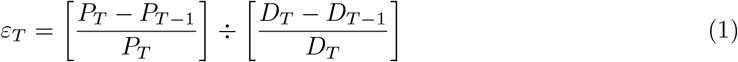

Rearranging the terms of the latter equation, we see that the acceleration index relates to the daily and average positivity rates, in the following way:

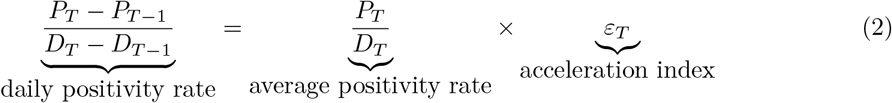

Equation (2) shows that the acceleration index is an elasticity, which is a concept widely used by economists since Marshall [19], to study the responsiveness of demand with respect to a change in price of a good. More precisely, the acceleration index is an elasticity that relates cumulated stocks (of cases to tests) over possibly extended periods if the epidemic lasts long. Mathematically speaking, such an elasticity measures the convexity of the relationship between cumulated cases and cumulated tests, using the non-local property that the linear approximation of any convex function provides a lower bound for that function at any point. In contrast, the second derivative is a local measure of convexity. Second, an important reason why we call such an elasticity an index of viral *acceleration* can be clarified using an analogy with linear body motions. Given that our analysis relies on data about cases and tests only, with the latter as units of measurement, one can think of the acceleration index as *the ratio between current and average viral speed*. With tests as the unit of measurement, the daily positivity rate *p*_*T*_ */d*_*T*_ becomes a measure of *current* viral speed at date *T*, that is, the fraction of tested people that turn out to be positive on that day. The average positivity rate *P*_*T*_ */D*_*T*_ at date *T* can be thought of as *average* viral speed, taken over the entire data sample. In Appendix B we illustrate through an example why we do not average over daily positivity rates in the usual way, but rather take the ratio of cumulated cases to cumulated tests. If then current viral speed is larger than average viral speed, we are in a situation of viral acceleration and the pandemic is on the loose. In that case, our acceleration index *ε*_*T*_ is larger than one, which means that increasing tests by 1% leads to *more than* 1% of new cases. An arguably legitimate goal of public health policy would therefore be to make sure that the acceleration index gets *smaller than one*, i.e. that current viral speed becomes smaller than average viral speed: this would indicate that the pandemic decelerates and becomes under control. Ideally, one would like to find ever fewer cases the more one tests. This reasoning also shows why it is not sufficient to look at positivity rates alone - they only indicate viral speed. What matters for public health is to understand whether speed becomes greater or smaller compared to its historical average as tests increase, which is what our acceleration index measures. In Appendix B, we give an example using exponential growth, for which closed-form solutions are derived and can be used to further illustrate the interpretation of the acceleration index as a unit-free elasticity that relates to how the current viral speed compares to its historical average.

Regarding the reproduction number, we make a rather general assumption, in accordance with the mathematical literature on epidemics, that the reproduction number is essentially a gross rate of growth and, as such, can be written at date *t* as:

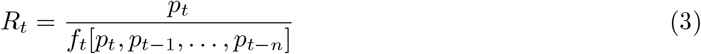

where *f*_*t*_ is a function of new cases from date *t* to date *t* − *n*, which can be thought of as the infectious potential, that is, the average number of people who have been infected at *t* and before, and who can infect people at *t*. The lag parameter *n* is related to infection duration. The assumed time dependence of *f*_*t*_ may capture many different phenomena that influence the number of cases, including for example health policy decisions but also the emergence of new strains of the virus. However, one specific factor that we have in mind here is the observation that the amount of performed tests is time-varying and so will be cases. Specifications for *f*_*t*_ have been used in the literature. We focus in this paper on a specific method, captured by Fraser’s [13] equation (9) - see page 3 of his paper - which defines the time-varying effective reproduction number as follows:

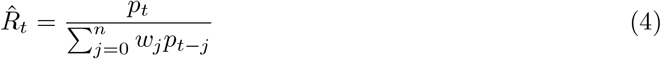

where the weights *w*’s capture the generation time distribution, with 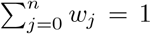. This means that the time-independent function 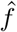 that follows from the denominator in equation (3) is, in that case, assumed to be *linear in the number of cases p* (which does not imply that *p*_*t*_ is linear in time of course). Note that such an assumption implies that, *given the reproduction number* 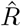, the dynamics of new cases follow an auto-regressive process *AR*(*n*). Although for the sake of presentation we focus on this specific method to estimate the effective reproduction number, our analysis extends to possible alternatives.

Even though it might go unnoticed at first sight, we should stress that a major difference between the rather general definition of *R* in equation (3) and the specific definition of 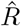 in equation (4) is that the function *f*_*t*_ implicitly depends on calendar time *t*. That is, what the latter takes account of is simply the number of cases detected, but not the fact that those cases will depend on the diagnostic effort or number of tests that has been realized. The fact that the diagnostics dimension is largely ignored in the literature about SIR-type models surfaces, for instance, in Wallinga and Lipsitch [24], who relate the epidemic growth rate to incidence and generation time interval only. It seems reasonable to assume that infectious and emerging diseases involve a diversity of pathogens, which require a variety of technologies to diagnose. In the context of COVID-19, PCR and antigen testing is of course key. This difference in accounting for cases turns out to be important to understand the connection between the acceleration index and the reproduction number, as we now show.

Using equations (2) and (3), we can relate our acceleration index and the reproduction number in the following way, at end date *T*:

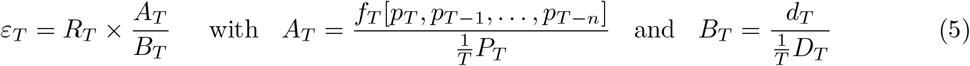

Equation (5) shows that the *ratio* between the acceleration index and the basic reproduction number can be itself decomposed into a ratio. The numerator of this latter ratio, *A*, can be thought of as the current infectivity intensity, that is, the ratio of the average number of primary cases up to period *T* who can originate infections in *T* as a fraction of the historical average of the number of persons who have been infected since the outset of the pandemic. The denominator, *B*, on the other hand, represents the number of tests in period *T* compared to its historical average up to *T*, that is, the current test intensity. To sum up, the ratio of the acceleration index to the reproduction number is, in any period, the ratio of the infectivity intensity to the test intensity.

From Equation (5) we see that both indicators are equal at all dates *t*, that is, *ε*_*t*_ = *R*_*t*_, if and only if:

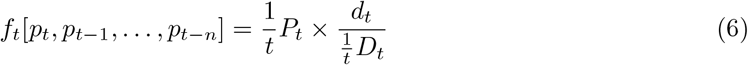

Equation (6) is very important to conceptualize the core idea of this paper: in order to properly control for the (time-varying) volume of tests/diagnostics, one needs to use the appropriate function *f*_*t*_, that is, one which depends on calendar time because tests do. Said differently, the function *f*_*t*_ should be specified in such a way that it takes account of the fact that cases are produced by tests or any other diagnostics. The linear form with no time dependence which appears in the denominator of equation (4) is therefore problematic, as it assumes away tests which are however key to measure the pandemic’s dynamics. In this sense, the acceleration index *ε* nests the basic reproduction number *R*: if the function *f*_*t*_ is specified as in equation (6), *R* is equivalent to *ε* as it takes account of testing; in any other case, *ε* is more general than *R*, as defined for example in equation (4) that takes account of cases only.

So as to elaborate more on why the acceleration index nests the reproduction number, in the sense that the former is a test-adjusted version of the latter, let us consider two hypothetical cases. The first case obtains when daily tests are constant at all dates, which implies that 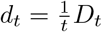, i.e. *B*_*t*_ = 1, and that equation (6) now reads as 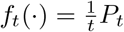. This means that in the case of time-invariant tests, the acceleration index and the reproduction number coincide if and only if the numerator of the infectivity intensity *A*_*t*_ is equal to the time average of all cases since the initialization date. This contrasts with the denominator in the expression of 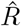 in equation (3): it defines a time-independent function 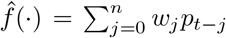 as a moving average of the number of cases over a time window of length *n* + 1, which implies that equation (6) is then violated. In the rather specific configuration such that tests are constant over time, one can see that the acceleration index and the reproduction number, as defined in (3), might differ because of the time window over which cases are included in the definition of the indicator of viral spread. More precisely, 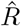 assumes a time lag that relates to the observed generation time of the disease and it is defined as the ratio of current cases over a weighted moving average. Since the latter tracks more closely the trend in the number of cases, 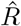 captures the proportional deviations from that short-run trend. In contrast, *ε* relies on the entire history of the pandemic since the number of cases is divided by a much smoother and longer-term trend (technically, the cumulative moving average), thus capturing shorter-term trends. The latter aspect is arguably relevant for public health decisions, due to possible path dependence and regime shifts related, for example, to mitigation policies and other changing behaviors. In addition, cases rising fast might make it more probable that new strains of the virus emerge(d) and revive the pandemic, thus creating a positive feedback loop. In addition, another benefit of de-trending the daily positive rate by the average positivity rate is to make *ε* a unit-free measure of viral acceleration that is useful to compare groups (see for example Baunez et al. [7] on vaccine effectiveness).

A more realistic configuration in view of the COVID-19 pandemic, however, is when tests do vary over time. Suppose they do but that, unrealistically, the daily number of newly found cases is now constant over time. In that case, 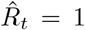 and the associated *Â*_*t*_ = 1. In such a situation, equation (6) is again violated because 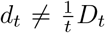, hence *B*_*t*_ ≠ 1. Such a violation signals not only that the acceleration index and reproduction number do not coincide, but also that the latter is not an accurate indicator of viral spread when tests vary over time but cases hypothetically do not. Either the test intensity is larger than one, meaning that the current level of tests exceeds its historical average so that 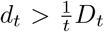 (hence *B*_*t*_ *>* 1). The reproduction number then overestimates viral acceleration compared to the acceleration index because, while new cases are still constant, current tests are above their historical average. In that case, the acceleration index is smaller than one and signals *deceleration* of viral spread, despite cases being constant over time. Or the test intensity is smaller than one (i.e. *B*_*t*_*<* 1), so that the reproduction number underestimates viral spread acceleration because current tests, while being below their historical average, still detect the same number of cases. The acceleration index is then larger than one, indicating indeed acceleration of viral spread. Such benchmark cases shed light on the reason why the reproduction number needs to be adjusted to take into account tests when they are time-varying.

A schematic example to help visualize the latter case is presented in Figure 1, assuming constant population size for simplicity. Suppose that the numbers of both tests and positive cases have been constant prior to date *t* and contrast the alternative outcomes at *t* + 1 (scenarii 1 and 2). At each date, a square represents total population and each dot represents one individual. Red areas include individuals who have been tested, among which green (respectively red) dots represent positive (respectively negative) individuals, while the complementary grey areas include untested individuals with unknown health status. From the situation at date *t* depicted in the left panel, two exclusive scenarii originate at *t* + 1, depending on whether the number of tests goes down (upper right, scenario 1) or up (bottom right, scenario 2), while the number of positive cases stay constant across scenarii, between *t* and *t* + 1. Note that using only the number of positive cases (hence ignoring the number of tests) leads to the conclusion that the epidemic situation has not changed since the (two-period) reproduction number 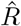 equals one at both dates. In other words, the epidemic situation neither worsens nor improves. However, comparing scenarii 1 and 2 in Figure 1 clearly reveals that the two alternative dynamics differ sharply: in the upper right panel, the number of tests goes down between *t* and *t* + 1, so that an equal number of cases is detected with a *smaller* number of tests, indicating an *accelerating* epidemic; in contrast, the lower right panel depicts a situation in which the number of tests increases markedly while an equal number of cases still materializes, indicating now a *decelerating* epidemic.

**Figure 1:**
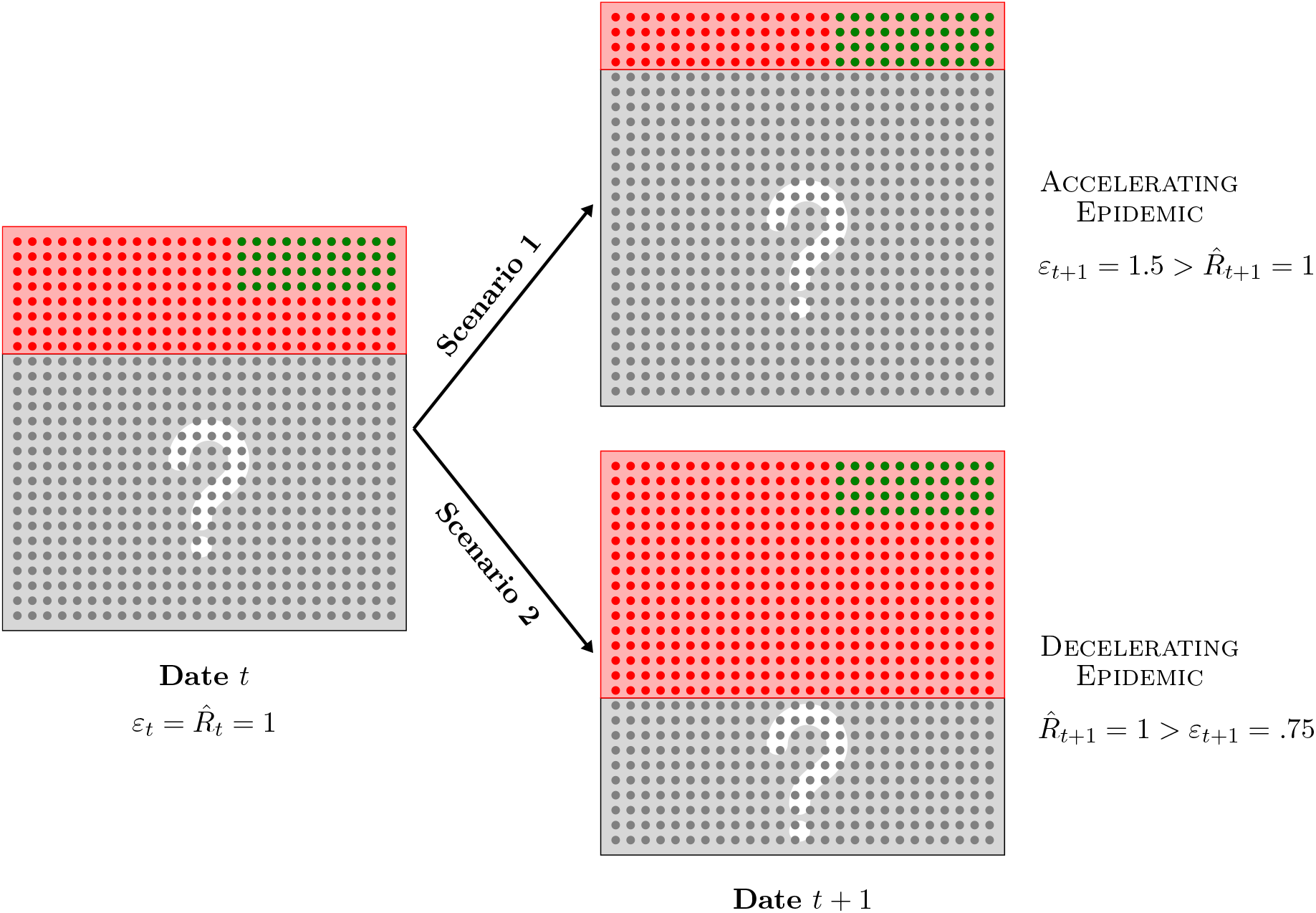
Schematic two-period example with both population and number of positive cases that stay constant over time. At each date, a dot represents one individual and red areas include individuals who have been tested, among which green (respectively red) dots represent positive (respectively negative) cases, while complementary grey areas depict untested individuals with unknown health status. From the situation at date *t* depicted in the left panel, two exclusive scenarii originate at *t* + 1 depending on whether the number of tests goes down (upper right) or up (bottom right), while the number of positive cases stay constant in each scenario, between *t* and *t* + 1.

In addition, it is perhaps instructive to go through the logic that delivers the magnitudes for the reproduction number 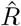 and acceleration index *ε* in Figure 1. Both equal one at date *t*, assuming again for simplicity an identical situation prior to that. Since cases do not changer over time, 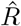 does not either and stays equal to one at both dates. In addition, it follows that a half of total cases - cumulated over the two periods - is detected at *t* + 1 equally in both scenarii. However the contribution to cumulated tests at *t* + 1 is not the same in both cases, compared to *t*. In scenario 1, tests are *halved* so that the contribution to cumulated tests at *t* + 1 is only 1*/*3 - that is, (1*/*2) ÷ (1 + 1*/*2). As a consequence, *ε*_*t*+1_ = (1*/*2) ÷ (1*/*3) = 1.5 in scenario 1: since *as much as* a half of total cases cumulated over the two periods is detected using *only* a third of cumulated tests, the epidemic is accelerating in period *t* + 1, as signalled by the property that 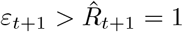 then. In contrast, the number of tests *doubles* at *t* + 1, in scenario 2, so that the contribution to cumulated tests is now 2*/*3 - that is, 2 ÷ (1 + 2). It follows that *ε*_*t*+1_ = (1*/*2) ÷ (2*/*3) = 0.75: the epidemic is decelerating since *only* a half of cumulated cases is detected using *as much as* as two thirds of cumulated tests, implying that 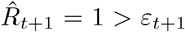. This example further illustrates why taking into account time-varying tests makes for a more accurate measure of epidemic acceleration. Note also that while the simple example in Figure 1 may give the impression that the positivity rate suffices to capture the epidemic dynamics, it is worth reiterating that it is a measure of speed which is not unit-free, as opposed to the acceleration index which is indeed a unit-free measure of the extent to which viral spread accelerates. Finally, incidence rates are not really useful to capture viral dynamics when population is constant, as in the example, or, more realistically, changing slowly.

Two general observations follow the above description of benchmark cases. First, when test intensity is larger (respectively smaller) than infectivity intensity, the reproduction number tends to over-estimate (respectively underestimate) viral acceleration compared to the acceleration index. This implies that the reproduction number must be test-adjusted if it is to serve well as an accurate enough indicator of viral spread that guides public health policies. Second, following the logic of the first hypothetical case outlined above, one might envision also versions of the test-adjusted reproduction number that would divide the expression in (3) by a similarly defined growth rate of tests, over a rolling window. For instance, a short-term test-adjusted reproduction number could be alternatively defined as:

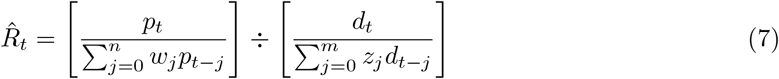

given the lag parameter *m* and 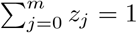. The expression in (7) is a simple test-adjusted version of (4). It would be interesting to investigate the properties of such a test-adjusted reproduction number, defined over rolling windows, and compare it to the acceleration index. As underlined above, keeping all the past of the pandemic is important, for instance to compare groups and derive a unit-free measure of vaccine effectiveness (see Baunez et al. [6, 7]), and possibly to capture path dependence. Although not addressed in this paper, whether the new indicator that is defined by equation (7) may turn out to be useful for other purposes is an open question. Finally, although this is beyond the scope of this paper, which abstracts from specific models of epidemics, we would like to stress some unreported results from simulation exercises. In a SIR model augmented to include time-varying tests, and in which only tested individuals among infected ones are observed, we have performed simulations indicating when the acceleration index captures more accurately the epidemic peak and deceleration than the reproduction number with imperfect information (and not test-adjusted). Interestingly, this happens in particular when tests become progressively available in a way that might lag the unobserved epidemic peak. Although of course model-and-parameter-specific, such simulation results go in the direction of the model-free results in this paper that test-adjusted versions of the reproduction number, such as the acceleration index that we advocate, better track the dynamics of viral spread.

To go back and better grasp the relation between the more general reproduction number *R* (not to be confused with 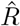) and *ε* as indicated in equation (5), a more theoretical analysis and its implications may be helpful. First of all, as it also becomes clear from equation (5), when *A* = *B* - that is when equation (6) holds - then *ε* = *R*. This basically means that if the test intensity *B* tracks the dynamics of the infectivity intensity *A*, there is enough testing to capture viral activity. In fact, seen from this perspective, we have a clear testing strategy: the daily tests *d*_*t*_ need to offset the assumed infectivity captured by function *f*_*t*_, and more specifically equation (6). The smaller the total number of cases, the easier it will be to match that testing requirement in particular through contact tracing. As total cases go up, contact tracing and sufficient testing may come to its structural and systemic limits. This in itself is a sign that additional health policies will need to be promoted.

In sum, if *A > B*, then *ε > R*, whilst when *A < B, ε < R*. In the former case, the infectivity intensity *A* of the pandemic cannot be matched by the test intensity *B* in place. That is, *R* does not give the appropriate picture of the infectiousness of the pandemic, in fact it underestimates it. To alleviate this bias, either testing would need to be increased, or viral spread would need to be cut by establishing policies that reduce contacts or a mixture of both. In any case, it shows that equation (3) that composes *R* does depend on more than past and current cases, because they themselves depend on tests and other factors that may favour or not transmissibility. Conversely, in the latter case, *R* will overestimate the speed of the pandemic if the test contribution *B* is greater than the infectivity contribution *A*. In such a situation, greater testing than underlying infectivity will necessarily find more cases, actually too many to reflect the correct transmissibility. To capture the correct picture, either testing would need to be reduced, which however seems counterproductive at least to the extent that testing is a way to look at the underlying viral dynamics, or the infectivity function *f*_*t*_ of equation (3) needs to be adapted to reflect reduced transmissibility.

In the next section, we apply the theoretical decomposition outlined above to capture how the reproduction number and acceleration index differ in the context of the current COVID-19 epidemic in France.

## 3 Results and Discussion

Before we turn to the application of the above analysis to French data, it might help to dissect a simple example showing in more details how and why the reproduction number and the acceleration index might differ. A thorough analysis of (5) requires structural assumptions, in particular to generate predictions about how both *ε, R* and their ratio move over time. In fact, the simple case of deterministic exponential growth, following Fraser’s [13] equation (12), comes in handy here. Time is assumed to be continuous, to ease derivation of results, and the number of cases grows exponentially over time, as usually assumed in epidemiological models, of SIR type and related for example. In such a case, the continuous-time analog of 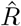 (not *R* now) defined in equation (4) is constant and any difference between 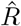 and *ε* is due to differences between *A* and *B*. For *ε*, we also have to introduce tests and we assume that they also grow exponentially.

Under those assumptions, we show in Appendix B that while the reproduction number is constant over time, the acceleration index is not, as it features different regimes depending on how the growth rate of daily cases compares with the growth rate of daily tests. For example, when the former is larger than the latter, the acceleration index first rises and then approaches a plateau, where it equals the ratio of growth rates, which is larger than 1 in that case. In contrast, the reproduction number stays constant over time. We can visualize this more easily in the simple setting of exponential growth (see Appendix B), but it also holds more generally that the difference between both indicators is essentially due to the fact that *while the acceleration index is the ratio of two growth rates, that of cases divided by that of tests, the reproduction number tracks only the former, thus ignoring the latter*. It is also for this reason that we say the acceleration index *ε* nests the basic reproduction number 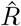, which is simply a special case of *ε*. Different configurations may in principle occur, therefore, over time, depending on how fast cases grow compared to the growth of tests.

To further illustrate what happens in the case of exponential growth outlined above and studied in more details in Appendix B, we now provide an illustration such that the growth rate of daily cases is twice as large as the growth rate of daily tests. Figure 2 illustrates how the acceleration index and the reproduction number, as well as the infectivity and test intensities, evolve over time in this particular example. In Figure 2, panel (*a*), we report the evolution over time of the time-varying acceleration index *ε*(*t*) and the constant reproduction number 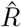 that follow from the numerical example. In Appendix B, we show that while 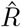 is constant, *ε* tends to the ratio of growth rates, which is equal to 2 in the example. As a consequence, a first regime with the reproduction number exceeding the acceleration number happens, followed by a second regime that features the reverse configuration. Not surprisingly, panel (*b*) in Figure 2 shows that the first regime materializes before day 9, when *B*(*t*) *> A*(*t*) - that is the test intensity exceeds the infectivity intensity - while the second regime is associated with *A*(*t*) *> B*(*t*) after day 9. Panel (*b*) reveals in particular that the plateau for the acceleration index that is featured in panel (*a*) comes from the fact that both *A*(*t*) and *B*(*t*) grow at the same rate, with the infectivity intensity exceeding the test intensity. Overall, therefore, the acceleration tracks the ratio of growth rates - which equals 2 in our example - while the reproduction number underestimates that ratio because it roughly reflects only its numerator.

**Figure 2:**
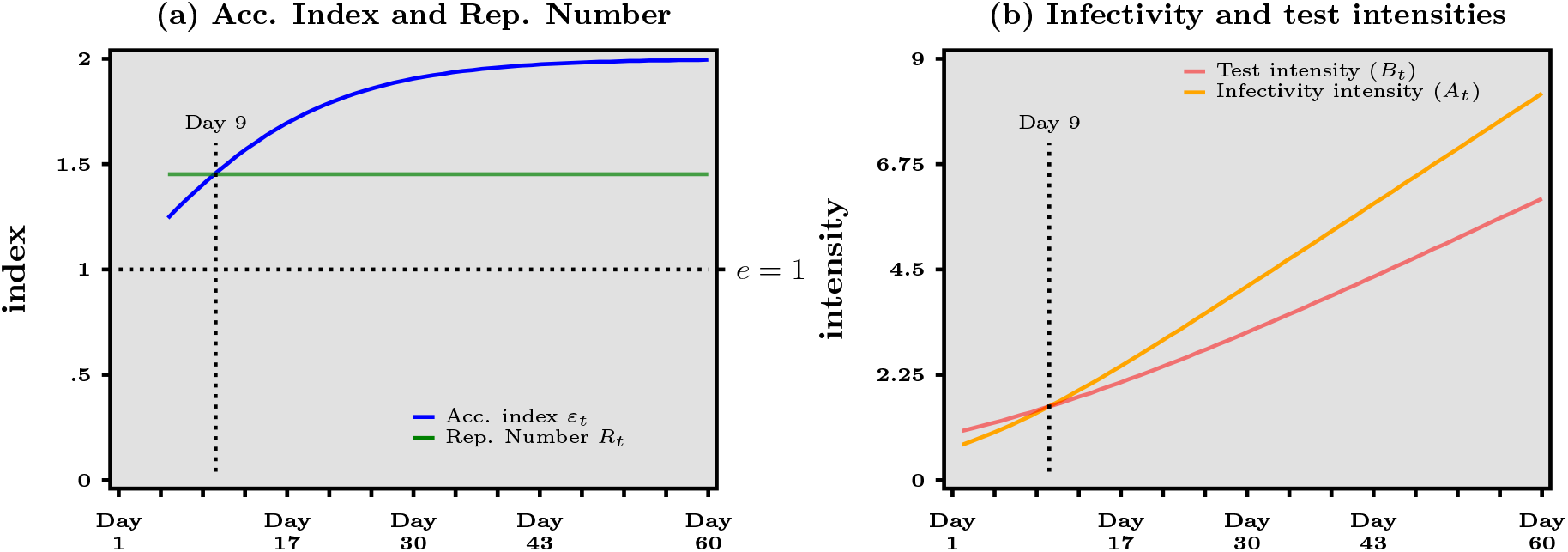
Numerical example for exponential growth of both daily cases and daily tests. Panel (a) Acceleration index (blue curve) and Reproduction number (green curve). Panel (*b*) Infectivity intensity (orange curve) and Test intensity (red curve)

Interestingly, we can derive from panel (*b*) in Figure 2 an operational tool to track, and possibly to control, in real time the difference between *ε* and 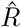. Figure 2 illustrates an acceleration epidemic phase, where positive cases grow faster than tests. As a consequence, test intensity *B*(*t*) eventually lags behind, infectivity intensity *A*(*t*). However, the implication that 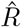 underestimates viral acceeration after day 9, compared to *ε* is not unescapable. In fact, one could aim at *controlling* the 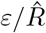 ratio: as data is updated in real-time, whenever it is observed that the infectivity intensity gets larger than the test intensity (that is, *A > B*), one should increase the latter by making sure that daily tests accelerate. Ideally, the test intensity should *track as closely as possible* the infectivity intensity, in order to optimize the accuracy of 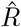 as an indicator of viral acceleration. Note that this goes in the same direction as the effort by public authorities to control viral spread by testing more and isolating the detected positive cases as much as possible. In that sense, to ensure that testing accelerates in the run up to an epidemic peak has two benefits: improving the epidemic situation, in so far as testing more contributes to the control of viral spread, *and* increasing the accuracy of the indicating tracking viral acceleration. Admittedly, tracking *ε* in real time is a more parsimonious way to attain accuracy, compared to tracking 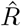 as well as *A* and *B* (or their ratio). Our decomposition shows formally that both approaches are equivalent though, and this result does not rely on exponential growth but holds more generally.

The conclusion that the reproduction number 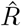 alone may poorly capture the dynamics of viral spread when tests vary over time, as expressed formally in equation (5), is not a mere theoretical curiosity. It strongly suggests possible pitfalls associated with its exclusive use in guiding and evaluating public health policies such as Non Pharmaceutical Interventions (NPI thereafter) in practice. Either *ε*, or 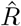 *together with* the ratio of infectivity and test intensities *A/B*, should be used to track viral acceleration. Casual reading of the literature reveals, however, that the reproduction number alone is widely assumed as the success metric to assess the effects of NPI (see e.g. Flaxman et al. [12] among many others). However, it should be clear by now that the conclusions thus derived should be considered with caution, at best, whenever tests are not accounted for. For example, Haug et al. [14] state that “In Fig. 4c, ‘enhancing testing capacity’ and ‘surveillance’ exhibit a negative impact (that is, an increase) on *R*_*t*_, presumably related to the fact that more testing allows for more cases to be identified.” Although increasing testing might indeed lead to an increasing reproduction number, it does not follow that such a NPI has an *adverse* effect on the epidemic, especially if tests are rising as much as during the March-April 2020 period considered by the latter authors in their study. Again, we cannot stress enough that whenever data about how many tests are performed is readily available (as in Haug et al. [14] but also for many other related studies), it should be used to measure as accurately as possible the dynamics of viral spread and adjust the reproduction number using property (6). Obviously, testing does not realistically capture all infected individuals unless all the population is tested each and everyday, but this observation should push policies towards testing as much as possible, not towards ignoring data about tests altogether. In addition, our analysis below clearly shows that completing the reproduction number with the positivity rate (that is, the ratio of positive cases to tests) is not a satisfactory answer either, as it might deliver conflicting evidence such as the former falling down to 1 and the latter shooting up.

Figure 3 summarizes our main empirical results obtained from data for France over the period that runs from May 13, 2020, to October 26, 2022. Note that data for France is available only for the period following the end of the first and longest lock-down, which extended from about mid-March to mid-May, 2020, and unfortunately not since the onset of the pandemic. More details about the input data and output variables used in the analysis of this section are gathered in Table 1 of Appendix A. In panel (*a*), we report both the acceleration index (blue curve) and the reproduction number (green curve) over time. The grey area represents the period before vaccination against COVID-19. Yellow areas depict the *second* and *third* lock-down periods. The yellow area ends more or less when the pink area starts, and this is when the first vaccination campaign begins around the end of year 2020 (see Table 2 of Appendix Afor precise dates). The acceleration index *ε* is computed using equation (1) while the reproduction number 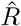 is computed from equation (4) with *n* = 7 and equal weights *w*. Note that the infection kernel could be adapted to account for sub-exponential growth as in Chowell et al. [9]. The lower spikes of the reproduction number 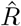 are due to much lower amount of testing during week-ends. This can clearly be seen in the panel (*d*) of Figure 3 that depicts the number of daily tests (in pink). We plot in all panels except (*c*) the raw variables rather any smoothed estimates in order to avoid any additional layer of interpretation.

**Table 1:** Summary of data description and computational processing

| Name | Variable | Computation |
| --- | --- | --- |
| Daily tests | $d_t$ | raw data from SI-DEP and Our World in Data |
| Daily positive cases | $p_t$ | raw data from SI-DEP and Our World in Data |
| Cumulated tests | $D_t$ | $\sum_{\tau=1}^t d_\tau$ |
| Cumulated cases | $P_t$ | $\sum_{\tau=1}^t p_\tau$ |
| Acceleration index | $\varepsilon_t$ | $\frac{p_t/d_t}{P_t/D_t}$ |
| Infectivity | $\hat{f}_t$ | $\frac{1}{n+1} \sum_{s=0}^n p_{t-s}$ |
| Reproduction number | $\hat{R}_t$ | $p_t/\hat{f}_t$ |
| Infectivity intensity | $A_t$ | $\hat{f}_t/(\frac{1}{t} P_t)$ |
| Test intensity | $B_t$ | $d_t/(\frac{1}{t} D_t)$ |

**Table 2:** Dates for main non-pharmaceutical interventions in France

| Name | Date begins | Date ends |
| --- | --- | --- |
| First Lock-down | Mar 17, 2020 | May 11, 2020 |
| Curfews | Dec 15, 2020 | Jun 20, 2021 |
| Second Lock-down | Oct 30, 2020 | Dec 15, 2020 |
| Third Lock-down | Apr 3, 2021 | May 2, 2021 |
| Vaccination | Dec 27, 2020 | still ongoing |

**Figure 3:**
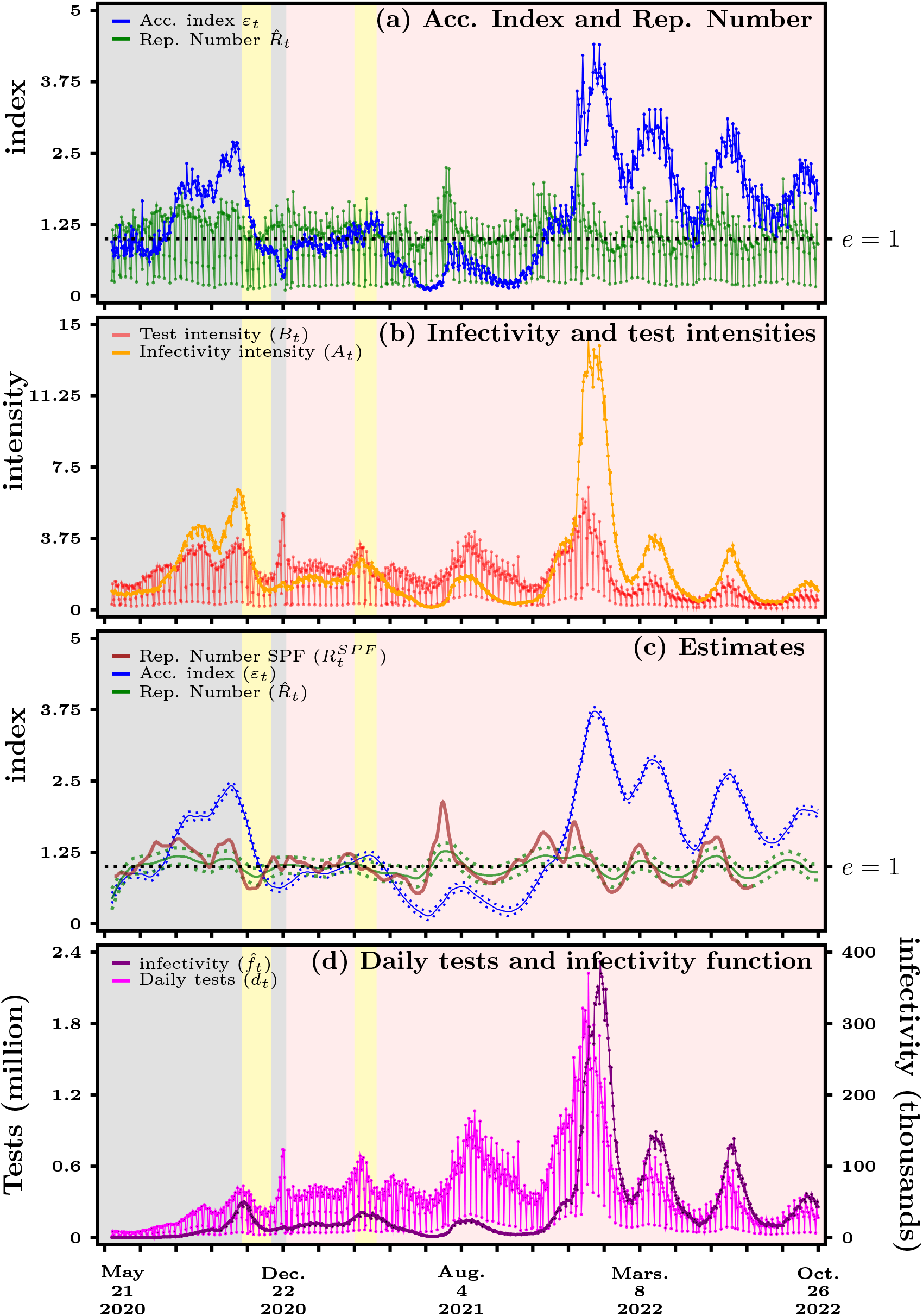
France - Panel (*a*) Acceleration index (blue curve) vs reproduction number (green curve). Panel (*b*) Infectivity intensity (orange curve) vs test intensity (red curve). Panel (*c*) Kernel estimates with confidence bands (dashed lines). Panel (*d*) Daily tests (purple line) and infectivity function (black curve). Yellow areas depict the second and third lock-down periods while pink area starts with vaccination. Source: Agence Santé Publique France and authors’ computations

In panel (*c*) of Figure 3, we present local polynomial regressions for 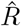 and *ε* of panel (*a*), that use the Savitzky-Golay filter also known as a locally estimated scatter-plot smoothing method in modern statistics (see Cleveland and Devlin [8]). The blue line is again our acceleration index. In red, we depict the reproduction number published by Santé Publique France, whereas the green line represents our own estimation of the reproduction number. As can be noted, the reproduction number estimated by Santé Publique France does not fall far outside the confidence bands of our own estimate which are the dotted green lines. Most importantly, both estimates of the reproduction number cross 1 at about the same dates. Even though Santé Publique France refers to the “Cori method”, we have not found public information about the precise weights attached to past values for the number of cases in computing infectivity. In addition, as can be seen from the confidence bands, the acceleration index is estimated more precisely than the reproduction number, because we take account of variations of tests and thus cases due to the week-end effect. In effect, being a ratio of growth rates (that of cases over that of tests), the acceleration index turns out to be smoother than the reproduction number (which is closer to the growth rate of cases and thus drops sharply over week-ends).

Let us now center the discussion around panel (*c*) of Figure 3 and focus first on the period preceding vaccination, which is represented by the grey area. We see that right after the end of the first lockdown, both indicators are hovering below 1, with 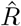 being slightly above *ε*. We concentrate here on the green line, i.e. our estimation of *R*, since the estimate by Santé Publique France is no longer included in the public data-set since August 12, 2022. Our estimated 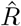 then rises quickly to a higher plateau in the first half of July 2020 to indicate greater transmissibility, and stays at a level of about 1.2 until mid-August 2020. At that same time *ε* first remains put at a level smaller than 1 and becomes greater than 1 a few days later, effectively crossing 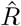 at the beginning of August and accelerating all along until about mid-August 2020.

The difference in dynamics of both indicators for France can easily be explained by looking at panel (b) in Figure 3. Here, we report the two terms that appear in equation (5), that is, *A*, the infectivity intensity (orange curve), and *B*, the test intensity (red curve). The latter graph exhibits spikes, again due to the fact that much less tests, if any, are performed during week-ends. The test intensity follows a downward trend that simply reveals the fact that tests being done in a given period constitute, over time, a smaller and smaller fraction of the cumulated amount of diagnostics. What we see in panel (*b*) in particular is that before July 29, the test intensity *B* is greater than the infectivity intensity *A* that has, at first, also a downward trend. A greater testing rate implies that more cases will be found. This corresponds to the period when 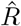 is greater than *ε*. But 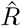 basically overestimates viral activity because it does not consider tests and focuses on cases only, while *ε* takes account of this because it looks at the ratio of both infectivity and test contributions. The opposite is true for the period after July 29, when the infectivity intensity *A* becomes greater than the test intensity *B*, hence *ε* greater than 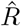. Despite growing daily tests until the end of August, as we can see in panel (*d*) of Figure 3, 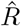 remains first at a plateau but then steadily declines until the second half of September.

Comparing *ε* with *R* over the summer of 2020 is even more striking. Soon after the acceleration index rises above the reproduction number, around August 5, 2020, both measures start to diverge and hence deliver opposite messages regarding the evolution of the pandemic. While *R* decreases from about 1.5 in August 15 to about 1 in September 22, *ε* goes up from approximately 1.7 in August 15 to 1.9 in August 25, and plateaus at the latter value until September 22. In other words, not accounting for time-varying tests over that period gives the impression that the pandemic situation improves, while on the contrary it is shown *to worsen* once we compare as we should the dynamics of the infectivity and test intensities. As shown in panel (b) in Figure 3, the period during which *R* goes down to reach 1 is also a period when the infectivity intensity *A* either rises faster or declines more slowly than the test intensity *B*: as a consequence, the acceleration index first rises and the plateaus around 1.9, indicating again a worsening of the pandemic. This in effect means that 1% of more of cumulated tests delivers 2% more of cumulated cases around September 22. A worsening of the pandemic indeed, which continues until the second lock-down depicted by the first and leftmost yellow area, with *ε* reaching about 2.5. While *R* also rises again from 1 after September 22, it remains true that looking *separately* at the reproduction number and at the positivity rate delivered conflicting messages about the pandemic resurgence, with the former indicating an improvement and the latter showing a worsening. In contrast, the acceleration index consistently indicated that the pandemic was still in an acceleration regime that at best stabilized before worsening again.

This clearly shows that 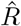 has been unable to represent viral acceleration basically during both summer months because, as we can see from panel (*b*), the infectivity intensity is rising more quickly than the test intensity. Therefore, 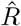 overlooks the testing dimension and only captures the number of cases, but those are undervalued given a lower test rate *B*. Even worse, testing then declines at the end of August while the infectivity function *f*, indicated in black in panel (*d*) starts going up. This affects 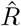 that declines to reach a level of about 1 by the end of September. This is in very stark contrast to *ε* that accelerates from early July onwards and then hovers at a plateau of about 2 up to end of September. It takes appropriately into account the relationship between the changing growth rates of testing and infectivity.

Both indicators go up again from the end of September onwards as testing rises again. But while 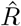 reaches a plateau again as the first curfew measures where put into place to cut transmission, *ε* further indicates acceleration. Both indicators then start declining when, at the end of October, the second lock-down was put into place. However, 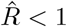 since the beginning of November, whilst at the same time, our indicator *ε* still shows an ongoing acceleration, although a reduced one with respect to the time before the lock-down. What we see very clearly from panel (*d*) is that lock-down coincides with a great reduction of testing. Obviously, lock-down is aimed at reducing contacts and thus viral spread. This will necessarily reduce cases and hence *R* declines. But if at the same time testing is reduced as well, which is the only way to get a clearer picture of the viral activity, this necessarily influences 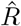 more dramatically and explains the under-evaluation of the viral spread than *ε* that continues to indicate acceleration. More specifically, *ε* indicates what happens to cases when we reduce testing by some percentage change. The fact that the percentage change of cases goes in the same direction as the percentage change of testing, i.e. that both decrease, is a good sign and indicates that lock-down measures have their effect. But looking at *ε* does not yet allow to give an all-clear such as *R* does. As a consequence, *ε* captures more accurately the considerable time variation of virus propagation than 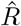.

Similar observations about the discrepancies between both indicators can be derived from the period that follows the first vaccination campaign in France, around the turn of year 2020, which is depicted by the pink area in panel (c) in Figure 3. In particular, the peak due to the Delta strain shows up as a rising acceleration index, with a local peak around August 4, 2021, which however stays in the deceleration regime with values much below 1. In contrast, both the estimate of Santé Publique France and our own estimate of the reproduction number take values much larger than 1 during the whole month of July 2021. Even more revealing is the period starting mid-November 2022, which saw the Omicron strain become progressively dominant. While the reproduction number computed by Santé Publique France goes *down* from about 1.6 on November 16 to about 1.1 on December 11, 2021, thus indicating an improvement, the acceleration index goes *up* from around 1 to 1.5 between those two dates, showing in contrast a return of the pandemic in the acceleration regime. Even more strikingly, the Omicron dominance period which begins at the end of December 2021 is associated with a continuous, albeit declining, worsening phase according to the acceleration index. In sharp contrast, the reproduction number shows alternating periods of exploding and dampening transmissibility, hovering around 1. To sum up, panel (c) in Figure 3 reveals that the year 2022 is associated with a significant bias of the reproduction number due to not accounting for time-tests (the magnitude of which can be seen in panel (*d*)). The origin of such a bias, we argue, is best understood using the decomposition between the test and infectivity intensities: as shown in panel (b) in Figure 3, over the year 2022 the infectivity intensity consistently wins the horse race against the test intensity, as the former stays larger than the latter. As a consequence, the reproduction number significantly underestimates the acceleration of viral spread, which turns out not to have been reduced by vaccines if one compares the grey and pink areas.

To make clear that the discrepancy between the reproduction number and the acceleration index shown in Figure 3 is not specific to France, we report in Appendix C a similar decomposition for five other countries over the pre-vaccination period. Such a comparison reveals that the reproduction number might either underestimate or overestimate viral acceleration, depending on the country and the time period, due to not correcting for the time-varying amount of testing. This readily suggests that this issue might be highly relevant in many other countries as well, where public health authorities are also in dire need of accurate indicators to track epidemics. As such, this observation should also preoccupy international bodies that design cooperation strategies to fight pandemics, including of course the World Health Organization and other regional agencies.

There is a number of interesting observations that can be drawn from the different experiences in France and those other 5 countries, which, however, should be independently evaluated in later research. First, in periods where the infectivity intensity *A* is greater than the testing intensity *B*, the acceleration index is larger than the reproduction number. This means that in those acceleration phases, the testing strategy is lagging behind the acceleration of viral propagation, so that the reproduction number is consistently underestimating acceleration of viral spread. This is the empirical confirmation of what we have already seen in our hypothetical numerical example for exponential growth in Figure 2. Second, increasing daily testing alone is not sufficient to track viral spread as we can note from observing the differences in the respective panels (*b*) and (*d*). In particular panels (*b*) and (*d*) for Austria and UK show that despite a high and increasing amount of daily testing, their test intensity remains more or less stable and does not match closely the infectivity intensity. However, in South Korea, but also in Argentina, test intensity does track rather closely the infectivity intensity and therefore, the gap between our acceleration index and the reproduction number is much smaller. It would be interesting to know how health authorities decided on their testing strategies in those two latter countries. Third, the reproduction number, which is agnostic about tests, is a poor indicator to guide efficient testing strategies. However, what we may be able to deduce from those empirical examples is that as soon as the acceleration index turns out to be greater than 1, there is a clear sign that diagnostic effort need to accelerate, i.e. that the test intensity need to be increased. It seems likely that in such a case, testing may be not only be used as a learning instrument about viral dynamics, but also as an instrument to guide public health policies and combat that dynamics, possibly in addition to other health policy measures. Said differently, the response to an indication of viral spread acceleration (that is, acceleration index larger than 1) must also be an acceleration in an instrument to offset that trend (a rising test intensity).

To further illustrate the differences between 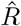 and *ε* and their consequences for public health decisions, we consider Figure 4. We use a background map that is in the public domain and can be downloaded via https://commons.wikimedia.org/wiki/File:D%C3%A9partements_de_France-simple. svg. Figure 4 is then constructed by incorporating our own data into this background map and gives an overview of how the second lock-down, which started on October 30, 2020, has contributed to reduce virus circulation across French départements. Looking at the two bottom maps shows that the acceleration index has been reduced everywhere during the period from October 30 to November 19. While a similar improvement is indicated by the reproduction number, as one concludes from the top maps, that measure of virus spread tells an altogether different story. Three weeks after the beginning of the second lock-down in France, the acceleration index suggests that the acceleration regime still prevails, except for 6 départements which happen to be the happy few, but with values still close to unity. However, one concludes rather wrongly from the reproduction number that, at the same date, deceleration is underway in most départements in green. In view of the discussion about the 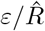 ratio in Section 3, this comes as no surprise since the reproduction number under-estimates virus circulation, due to the fact that it does not take tests into account.

**Figure 4:**
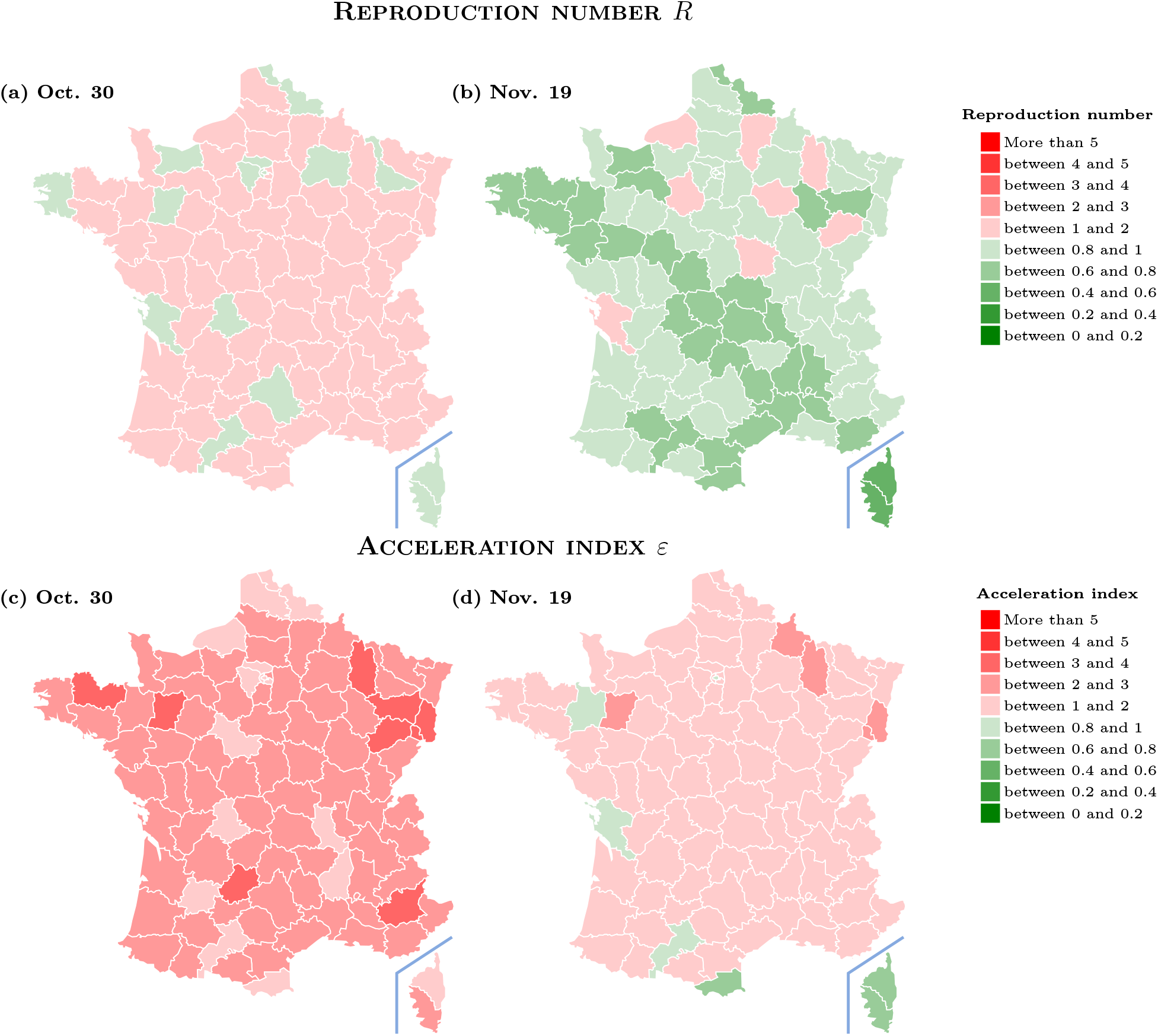
Acceleration index and reproduction number for French départements at different dates. October 30 dates the start of the second lock-down in France. Data source: Agence Santé Publique France and authors’ computations.

To sum up, two main differences between the reproduction number and the index appear in panel (*c*). The first being that 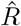 crosses unity earlier than the acceleration index, which starts to increase around July 6. This is most likely due to the infectivity rate reaching its lowest point around that date (as seen in panel (*b*)). Passing that date, the infectivity rate begins to increase, while the test rate continues downwards. This explains why the acceleration index could not start growing before July 6. Overall, therefore, 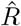 is larger than *ε* before August 5. The second difference is a sudden decrease of the reproduction number in the second half of September, while the acceleration index stays at a plateau. This can be explained by panel (*d*), in which we see a sharp plummet in the number of daily tests around that period. Less tests equals less detected cases, which 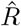 relies heavily on for its calculation. Seeing 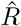 rising sharply from about 1 in early October is all the more surprising when seen in isolation. In contrast, the acceleration index, which accounts for variations of both cases and tests, consistently shows a succession of periods of steep rise followed by plateaus over the summer and until the second lock-down.

In practice, many public health agencies report (daily or weekly) positivity rates, to complement the information contained in the reproduction rate 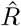. In light of the connection with *ε* that we have highlighted in this paper, both formally and empirically, we argue that the acceleration index is closer to a sufficient statistic that helps tracking the rapidly changing dynamics of any pandemic, because it explicitly takes into account the dynamics of diagnostics. All in all, the acceleration index is a test-adjusted reproduction number. In the context of COVID-19, diagnostics equal tests, but our claim is valid more generally when this is not the case. This means that the acceleration index can potentially be applied to any effort designed at detecting infected people, no matter what the pathogen agent triggering the infectious disease turns out to be. In real time, this is quite valuable, we believe, to guide health policies and to assess containment measures, especially in the context of a new pathogen appearing (such as SARS-Cov-2), with unforeseeable pandemic dynamics.

## 4 Conclusion

We show in this paper that the reproduction number is a special case of the acceleration index proposed in Baunez et al. [5]. While the former only considers the growth rate of cases, the latter measures variations of cases in relation to that of tests, and it does so in a unit-free manner since it is an elasticity. Most importantly, the acceleration index is a sufficient statistic of viral spread in the sense that it aggregates all the relevant information in a synthetic manner. In contrast, looking at pieces of information, like positivity or prevalence rates, *separately* might lead to the misleading conclusion that there is conflicting evidence about whether the epidemic worsens or improves. As such, a test-controlled reproduction number like the acceleration index should be part of any data dashboard to track an epidemic, and especially to guide public policy in the design of the most efficient methods to curb it. For example, we have shown in Baunez et al [5] that an accurate measure of virus circulation is a key input to feed algorithms that are designed to efficiently allocate the diagnostic effort across space.

The result that the reproduction number is, as a measure of viral spread, subject to a considerable bias is not specific either to France or to the period considered, as illustrated using data from five other countries. Such a comparison reveals that the reproduction number might either underestimate or underestimate viral acceleration, depending on the country and the time period, due to not correcting for the time-varying amount of testing. This readily suggests that this issue might be highly relevant in many other countries as well, where public health authorities are also in dire need of accurate indicators to track epidemics. As such, this observation should also preoccupy international bodies that design cooperation strategies to fight pandemics, including of course the World Health Organization and other regional agencies.

Relatedly, a key conclusion follows from our theoretical and empirical results. If public health authorities aim at measuring as accurately as possible viral acceleration, they have to rely on one of the following strategies: track in real time either the acceleration index alone, or a combination of the reproduction number together with the test and infectivity intensities. Although both strategies are formally equivalent, the latter is not only less parsimonious, it is also arguably more delicate to operate in practice since one would then like to control the bias that inevitably comes from time-varying tests. This is one of the main reasons why we argue in favor of using the acceleration index.

Such observations make the acceleration index a more parsimonious indicator to track a pandemic in real-time, as it is context-dependent: the acceleration index takes into account the effort to diagnose people who have been infected by the pathogen. In the case of COVID-19, diagnostics equal PCR (and other types of biological) tests, but this might not be the case for other diseases where diagnostics require even greater effort. However, our analysis makes a strong case for incorporating in any measure of pathogen circulation the observed effort to diagnose the agent that makes people sick.

Even though there is a variety of infectious (and emerging) diseases, with different pathogens and various ways to diagnose them, we claim that our conceptual approach is general enough to shed light, not only on the current pandemic, but also on any future ones which may come. In addition, our analysis extends to alternative methods to estimate the effective reproduction number, beyond the specific example stressed in this paper for the sake of presentation.

Finally, we would like to mention some limitations of our analysis. Some important issues, beyond the scope of our paper, have however been addressed by the literature. First, the fact that unaccounted cases arise when testing is not compulsory (see Pullano, Di Domenico, Sabbatini et al. [21] for France). Second, the coexistence of symptomatic and asymptotic cases during COVID-19 has led to additional statistical methods (see Khailaie et al. [18]). Another limitation of our acceleration index is that its accuracy depends positively on the amount of tests performed. For instance, detecting infected people who are asymptomatic but still potentially infectious requires an active policy and enough testing capacities. Even though the more tests the better in terms of how accurate the acceleration index is, should be included in the analysis that testing policy requires costly resources. From an economic standpoint, future research should also examine cost-efficiency of policies aims at mitigating epidemics, including using testing actively.

## Data Availability

Data used publicly available.

## A Data Description and Computational Processing

All the source data used in this study are available publicly from the web page “Données relatives aux résultats des tests virologiques COVID-19 SI-DEP” https://www.data.gouv.fr/fr/datasets/donnees-relatives-aux-resultats-des-tests-virologiques-covid-19/ for France and from Our World in Data via https://ourworldindata.org/coronavirus for the five other countries, from which we extract the raw daily data for both the number of tests performed on a given day and the associated positive cases detected the same day from those tests, over the period from May 13, 2020, to October 26, 2022, for France, and over the period from May 13 to November 19, 2020, for the five other countries. In Table 1, we provide both the definition of the input data and the computational processing for the output variables that are used in the empirical analysis of Section 3.

For convenience, we also report in Table 2 the beginning and end dates for the main non-pharmaceutical interventions against COVID-19 that have been implemented in France.

## B Exponential Growth: an Illustrative Example

We now illustrate the relationship between the acceleration index and the reproduction number when time is assumed, to ease derivation of results, to be continuous and when the number of cases grows exponentially over time, as usually assumed in epidemiological models, of SIR type and related for example. See for example Fraser’s [13] equation (12). Although typically ignored in the latter strand of literature, we introduce tests and we assume that they also grow exponentially. More formally, using the notation in the previous section, suppose that the number of cases per unit of time is denoted by *p*(*t*) = *αe*^*βt*^ while the number of tests per unit of time is *d*(*t*) = *γe*^*νt*^, where the growth rates *β* and *ν* are assumed to be positive for the sake of illustration. The reproduction number is then constant over time, as we now show when the infection kernel is uniform, that is, if the weights *w* are constant and equal to 1*/δ* to ensure that 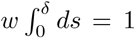. Note that it is not difficult to show that the constancy of 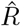 holds for other infection distributions as well. The analog of equation (4) is as follows:

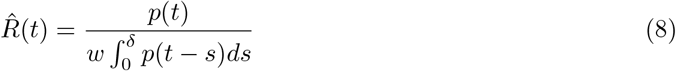

which implies that, given *p*(*t*) = *αe*^*βt*^, one has:

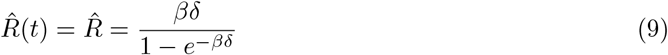

It is easily seen that the reproduction number is the growth rate of daily cases (adjusted for the delay *δ*) only, since it obviously does not take into account tests. In fact, 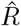 is the growth rate of the denominator in equation (8), that is, what we note 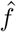 in the main text.

Cumulated cases and tests are then noted 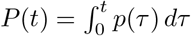 and 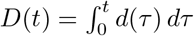, respectively. It is easy to derive, by straight integration, the expressions:

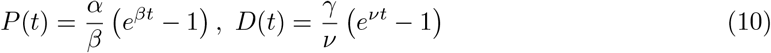

It follows that our acceleration index is given, as function of time, by:

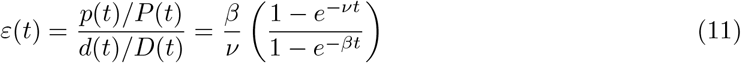

From equation (11), one concludes that the acceleration index, which is an elasticity that measures the responsiveness of cases to tests, is essentially the *ratio of the growth rate of cumulated cases divided by that of cumulated tests*, since *p*(*t*) = *dP* (*t*)*/dt* and *d*(*t*) = *dD*(*t*)*/dt*. In addition, *ε* tends to the ratio of the growth rate of daily cases to that of daily cases *β/ν*. This means that the acceleration index tracks that ratio over time and converges to it eventually.

It follows that three cases occur. When *β* = *ν*, that is, when both daily cases and daily tests grow at the exact same rate, then our acceleration index equals 1 at all dates. When the two growth rates differ, however, *ε*(*t*) converges, when *t* goes to infinity, to the ratio of growth rates *β/ν*, independently of the scale parameters *α* and *γ*. As an illustrative example, suppose that *β > ν*, so that positives grow faster than tests. Then the pattern of our acceleration index *ε*(*t*) over time will have two regimes: it first grows almost linearly and eventually reaches the upper bound *β/ν >* 1. Obviously, in that case both the daily positivity rate *p*(*t*)*/d*(*t*) and the average positivity *P* (*t*)*/D*(*t*) grow over time, and the latter quantity exceeds the former all the time so that acceleration prevails. The symmetric case when *β < ν* is easily adapted.

The property that the reproduction number is constant under our assumptions essentially means that the dynamics of the acceleration index is driven by that of *A*(*t*)*/B*(*t*), which we now decompose to help understand how 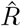 and *ε* compare over time. It follows from the definition of *A* and *B* in equation (5) that:

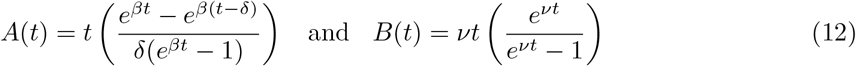

To further illustrate what happens in the theoretical case outlined above with *β > ν*, let us take a numerical example. Suppose that the growth rate of cases *β* equals 20% while the growth rate of tests *ν* = 10%. In addition suppose that the delay parameter *δ* = 4 so that the weight *w* = 1*/*4. Figure 2 illustrates how the acceleration index and the reproduction number, as well as the infectivity and test intensities, evolve over time in this particular example, with both functions *A*(*t*) and *B*(*t*) increasing with time *t*.

The exponential example is also useful to illustrate the formal relationship between viral speed and the acceleration index. To do that, let us now make an analogy with linear body motions to relate our indicators to speed and acceleration. If one think of the positivity rate as viral speed and define it as 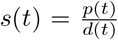 then straightforward computations give 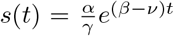. By analogy, viral acceleration could be defined as the derivative of viral speed, that is, 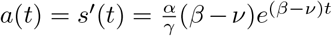. Not surprisingly, then, it turns out that the *sign* of *a*(*t*) indicates acceleration in the sense that a positive sign reveals acceleration (this happens when *β > ν* since speed goes up over time in that case) while a negative sign reveals deceleration (when *β < ν*).

However, a drawback of *a*(*t*) as a *measure* of acceleration is that it is scale-dependent since it depends on scale parameters *α* and *ν*, though their ratio. In other words, *a*(*t*) depends on the absolute level of speed, that is, of the positivity rate. On the contrary, the acceleration index given by Equation (11) does not and is unit-free, since it is an elasticity. Note that a possible, also unit-free, alternative would be to define acceleration as the semi-elasticity *a*(*t*)*/s*(*t*) = *β* − *ν*. However, since it relates the percentage change of positives case to the *absolute* change of tests, it is arguably less amenable to interpretation than the elasticity. It is easily shown that the semi-elasticity would be obtained if the average positivity rate would alternatively be defined as the average of the daily positivity rates up to date *T*, as opposed to the ratio of number of cumulated cases to the number of cumulated tests as in Equation (2) which leads to the acceleration index as an elasticity. Finally, note that the acceleration index equals one when viral speed stays constant over time.

## C Test-Controlling the Reproduction Number in Practice: Five Additional Countries

In this appendix, we provide estimates for both the acceleration index and the reproduction number, for Argentina, Austria, South Africa, South Korea and the United Kingdom, using the same method that is applied in Section 3 for France and over a pre-vaccination period (May 13 to November 19, 2020). The public data source for the five countries comes from Our World in Data (Ritchie et al. [22]). The resulting estimates, depicted in Figures 5 to 9 (that are hence directly comparable to Figure 3 for France), unambiguously reveal that the substantial bias due to time-varying tests, which is corrected by the acceleration index but not by the reproduction number, is not specific to France or to a particular period. In fact, the corresponding bias forces the reproduction number to either underestimate (Argentina over the whole period is a case in point) or overestimate viral acceleration, depending on the country and the period considered. This observation suggests that a similar bias should come as no surprise for other countries as well.

**Figure 5:**
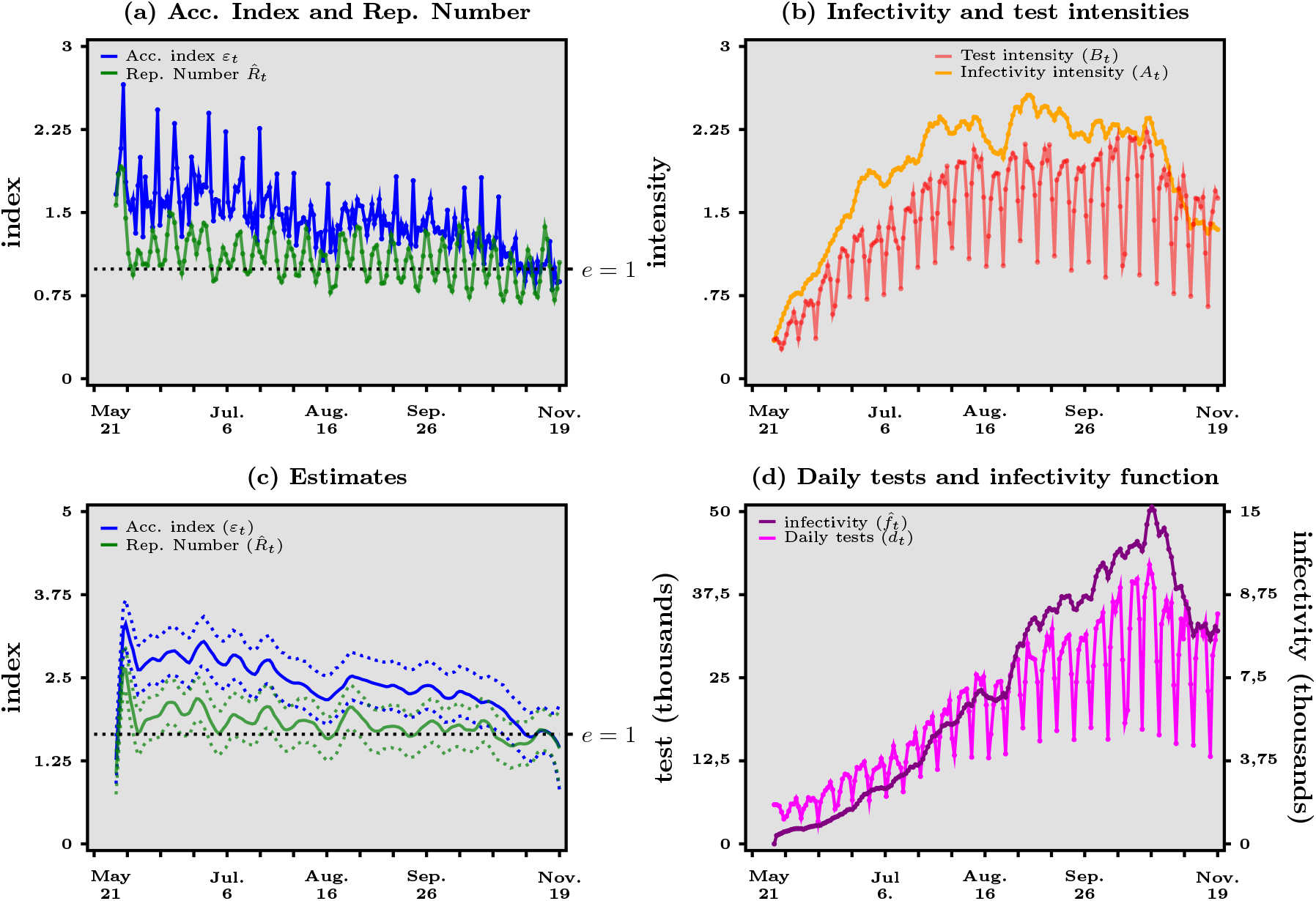
Argentina - Panel (*a*) Acceleration index (blue curve) vs reproduction number (green curve). Panel (*b*) Infectivity intensity (orange curve) vs test intensity (red curve). Panel (*c*) Kernel estimates with confidence bands (dashed lines). Panel (*d*) Daily tests (purple line) and infectivity function (black curve). Source: Our World in Data and authors’ computations.

**Figure 6:**
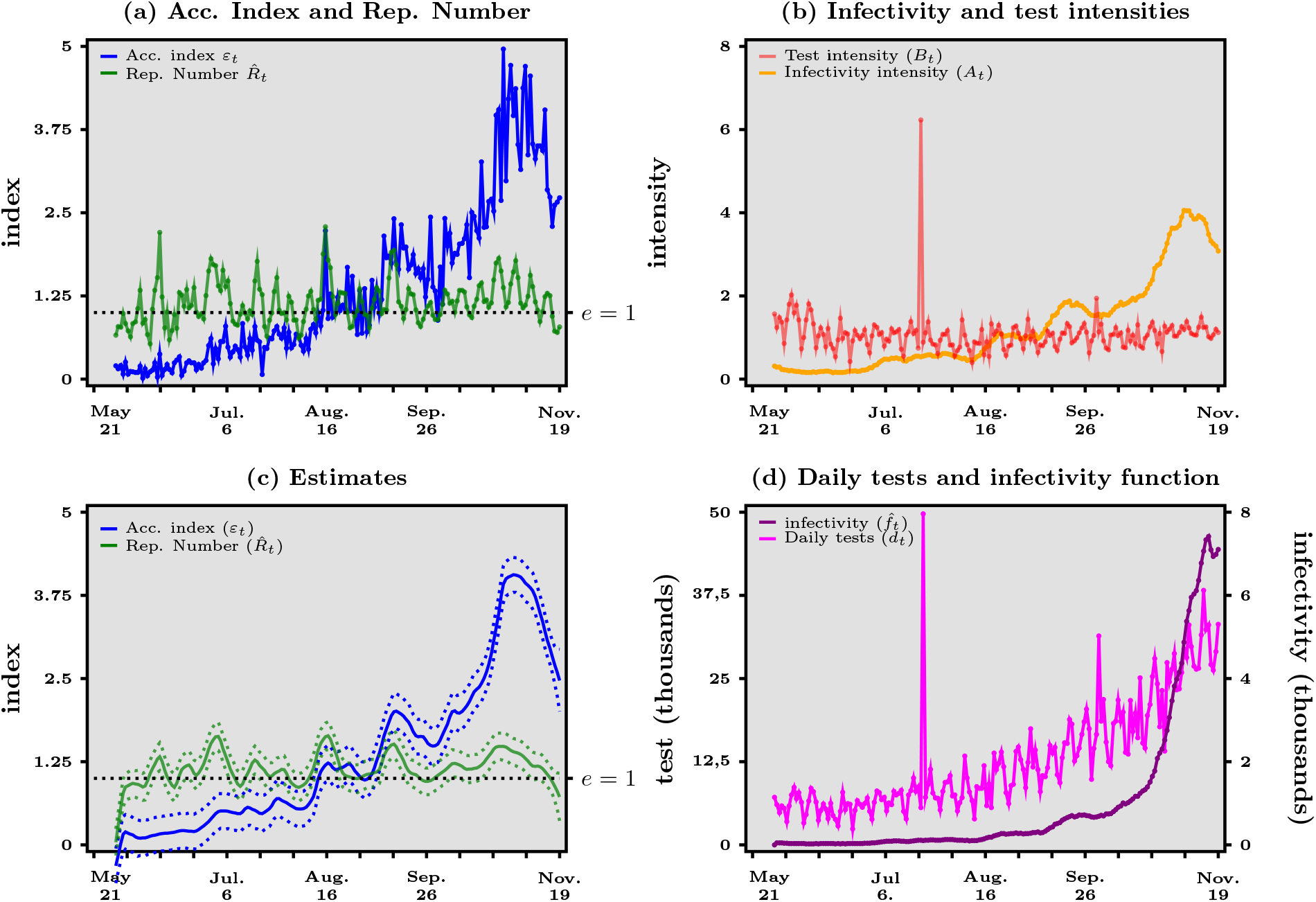
Austria - Panel (*a*) Acceleration index (blue curve) vs reproduction number (green curve). Panel (*b*) Infectivity intensity (orange curve) vs test intensity (red curve). Panel (*c*) Kernel estimates with confidence bands (dashed lines). Panel (*d*) Daily tests (purple line) and infectivity function (black curve). Source: Our World in Data and authors’ computations.

**Figure 7:**
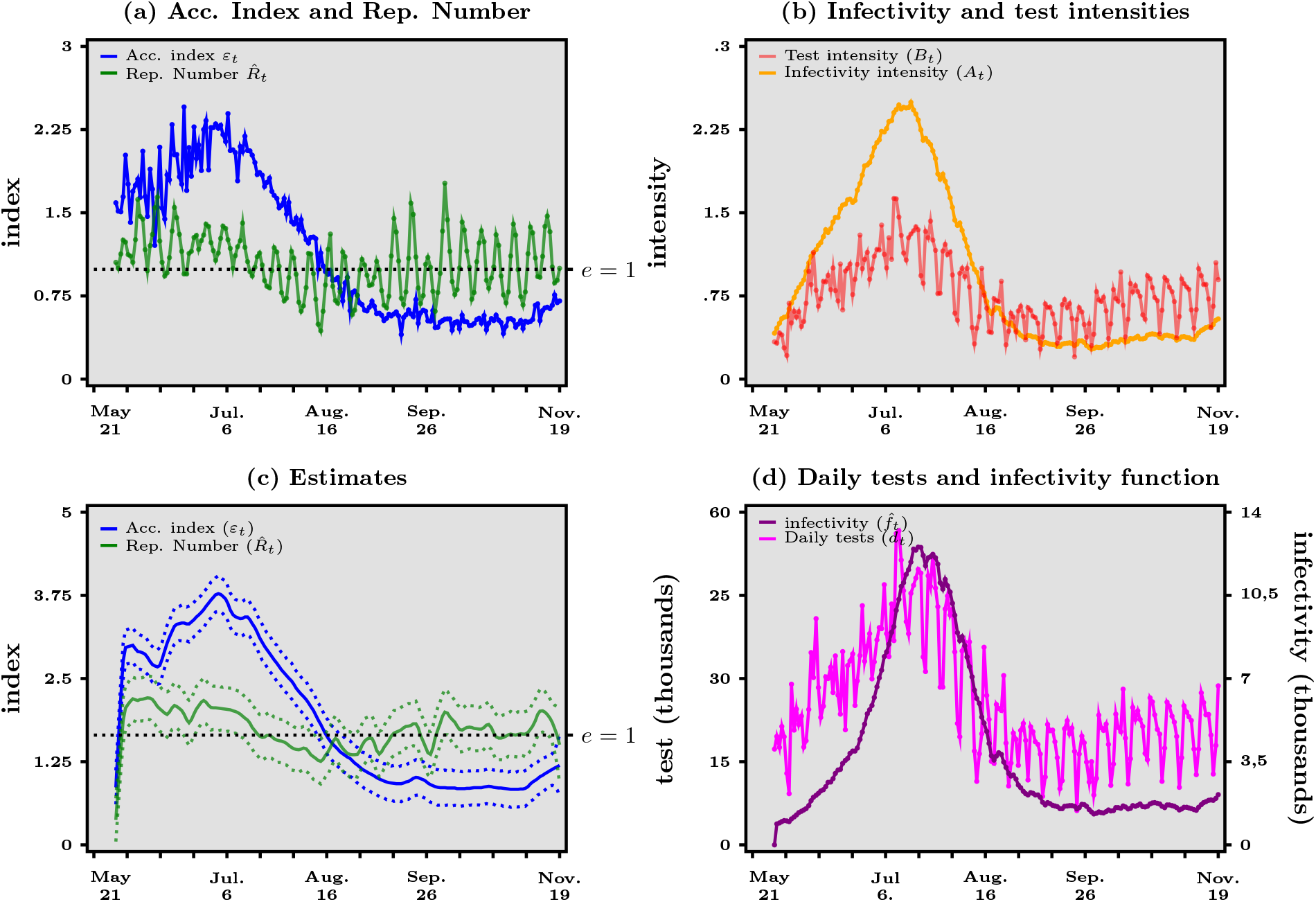
South Africa - Panel (*a*) Acceleration index (blue curve) vs reproduction number (green curve). Panel (*b*) Infectivity intensity (orange curve) vs test intensity (red curve). Panel (*c*) Kernel estimates with confidence bands (dashed lines). Panel (*d*) Daily tests (purple line) and infectivity function (black curve). Source: Our World in Data and authors’ computations.

**Figure 8:**
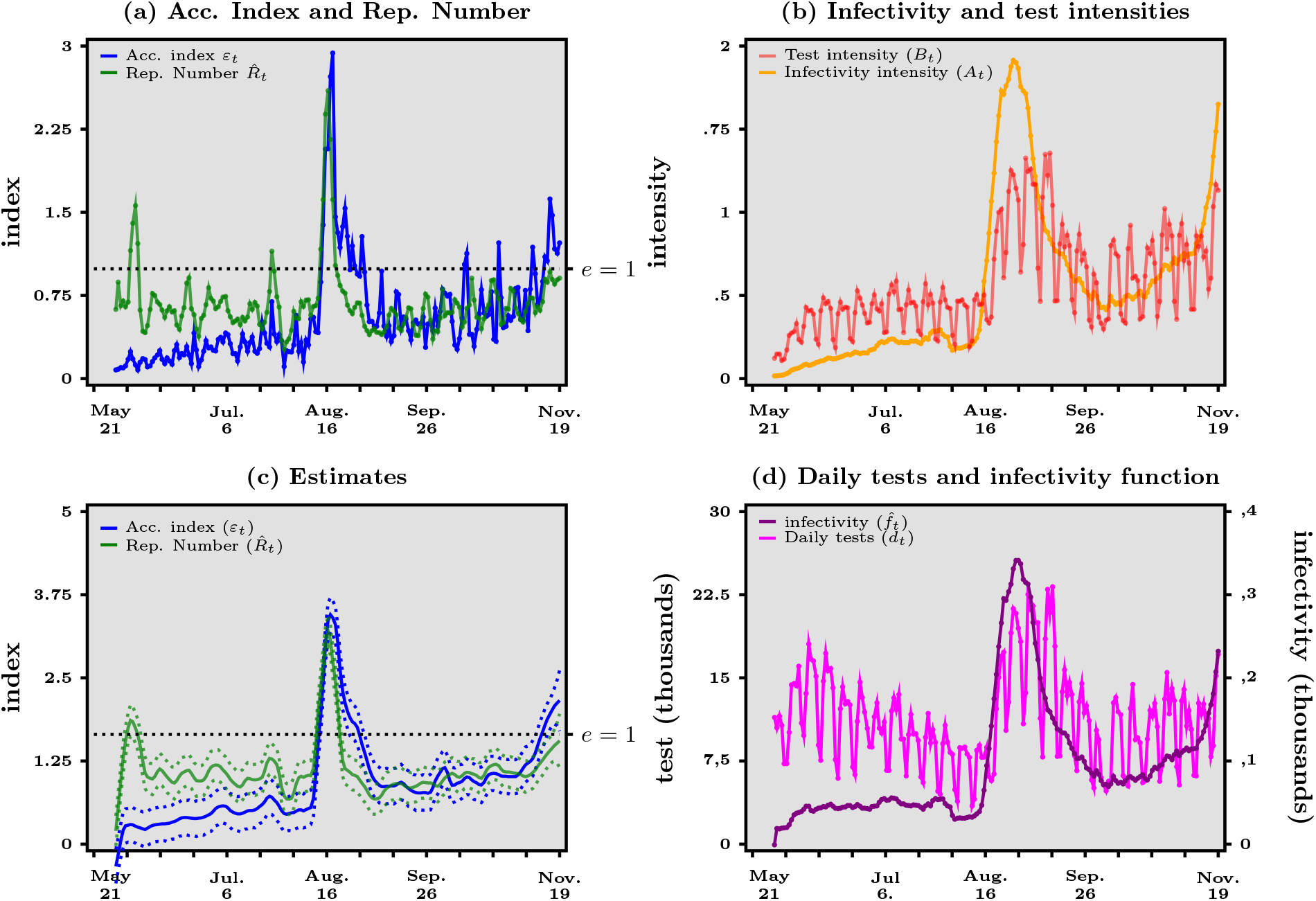
South Korea - Panel (*a*) Acceleration index (blue curve) vs reproduction number (green curve). Panel (*b*) Infectivity intensity (orange curve) vs test intensity (red curve). Panel (*c*) Kernel estimates with confidence bands (dashed lines). Panel (*d*) Daily tests (purple line) and infectivity function (black curve). Source: Our World in Data and authors’ computations. Testing data for South Korea were incomplete: five dates were missing. Missing values were completed by a linear interpolation.

**Figure 9:**
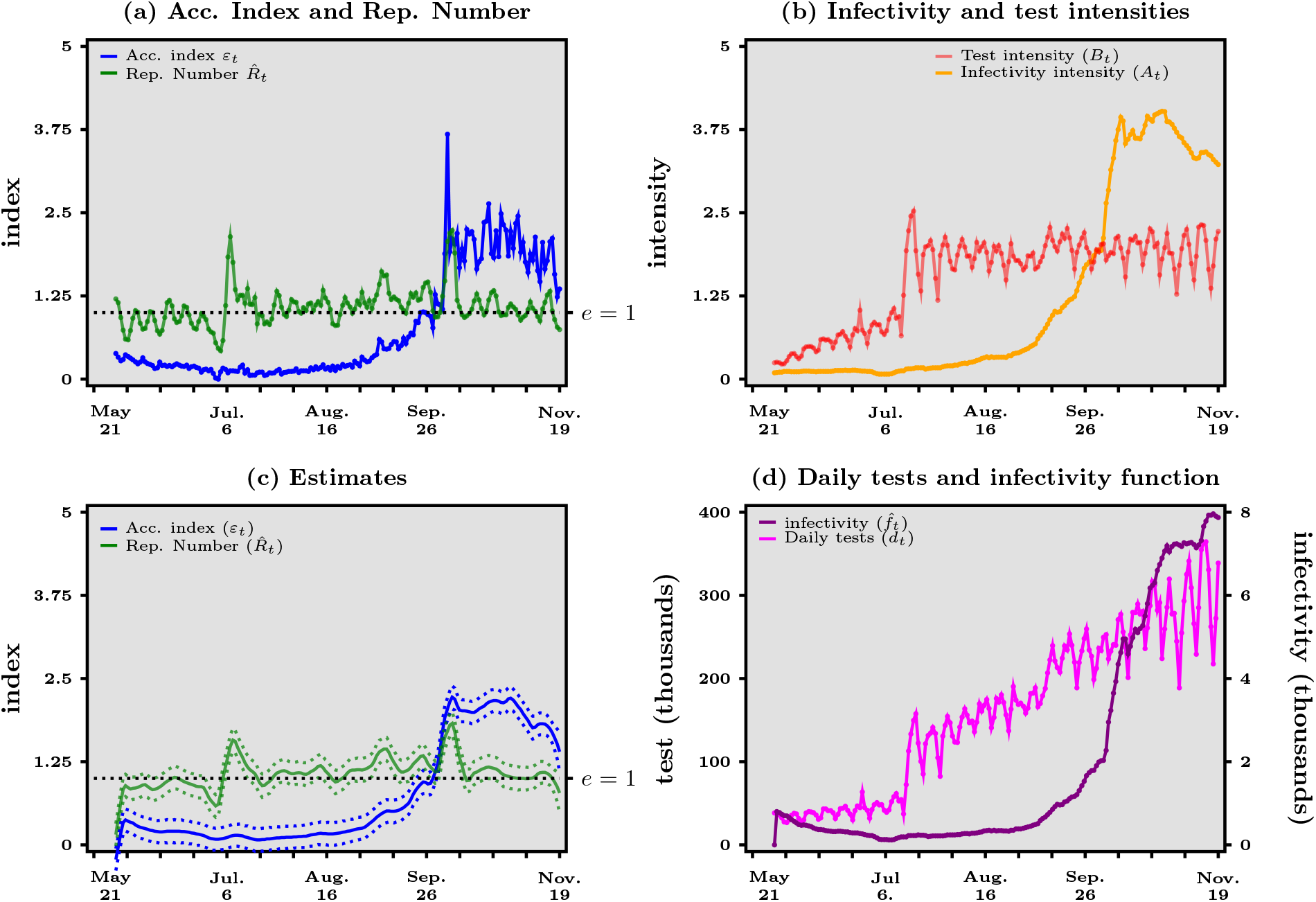
United Kingdom - Panel (*a*) Acceleration index (blue curve) vs reproduction number (green curve). Panel (*b*) Infectivity intensity (orange curve) vs test intensity (red curve). Panel (*c*) Kernel estimates with confidence bands (dashed lines). Panel (*d*) Daily tests (purple line) and infectivity function (black curve). Source: Our World in Data and authors’ computations.

## References

[1] Althaus C.L. (2014): Estimating the Reproduction Number of Ebola Virus (EVOB) during the 2014 Outbreak in West Africa. PLOS Currents, Sep 2:6. 4

[2] Bauch C.T., Lloyd-Smith J.O., Coffee M.P., Galvani A.P. (2005): Dynamically Modeling SARS and Other Newly Esmerging Respiratory Illnesses: Past, Present, and Future. Epidemiology, 16:791–801. 4

[3] Baunez C., Degoulet M., Luchini S., Pintus P., Teschl M. (2020): Sub-National Allocation of COVID-19 Tests: An Efficiency Criterion with an Application to Italian Regions. Covid Economics, 12:192–209.

[4] Baunez C., Degoulet M., Luchini S., Pintus P., Teschl M. (2020): An Early Assessment of Curfew and Second COVID-19 Lock-down on Virus Propagationin France. MedRχiv preprint 11.11.20230243 available at https://doi.org/10.1101/2020.11.11.20230243. 2, 5

[5] Baunez C., Degoulet M., Luchini S., Pintus P., Teschl M. (2021): Tracking the Dynamics and Allocating Tests for COVID-19 in Real-Time: an Acceleration Index with an Application to French Age Groups and Départements. PLoS ONE, June 1st, available at https://doi.org/10.1371/journal.pone.0252443. 2, 5, 21

[6] Baunez C., Degoulet M., Luchini S., Pintus P., Teschl M. (2021): COVID-19 Acceleration and Vaccine Status in France - Summer 2021. MedRχiv preprint 11.11.20230243 available at https://www.medrxiv.org/content/10.1101/2021.09.18.21263773v3. 2, 11

[7] Baunez C., Degoulet M., Luchini S., Pintus P., Teschl M. (2022): Vaccine Effectiveness against COVID-19 Infection from Real-Time Population Data in France. Forthcoming as MedRχiv preprint. 8, 11

[8] Cleveland, W.S., Devlin, S.J. (1988): Locally-Weighted Regression: An Approach to Regression Analysis by Local Fitting. Journal of the American Statistical Association. 83:596–610. 16

[9] Chowell G., Viboud C., Simonsen L., Moghadas S.M. (2016): Characterizing the Reproduction Number of Epidemics with Early Subexponential Growth Dynamics. Journal of the Royal Society Interface, 13(123):20160659. 16

[10] Cori A., Ferguson N.M., Fraser C., Cauchemez S. (2013): A New Framework and Software to Estimate Time-Varying Reproduction Numbers during Epidemics. American Journal of Epidemiology, 178:1505–1512. 2, 4

[11] Cori A., Donnelly C.A., Dorigatti I., Ferguson N.M., Fraser C., Garske T., Jombart T., Nedjati-Gilani G., Nouvellet P., Riley S., Van Kerkhove M.D., Mills H.L., Blake I.M. (2017): Key Data for Outbreak Evaluation: building on the Ebola Experience. Philos Trans R Soc Lond B Biol Sci. May 26; 372 (1721):20160371. 4

[12] Flaxman, S., Mishra, S., Gandy, A. et al. (2020): Estimating the effects of non-pharmaceutical interventions on COVID-19 in Europe. Nature 584:257–261. 14

[13] Fraser C. (2007). Estimating Individual and Household Reproduction Numbers in an Emerging Epidemic. PLoS One, 2:e758. 2, 4, 6, 12, 27

[14] Haug, N., Geyrhofer, L., Londei, A. et al. (2020): Ranking the effectiveness of worldwide COVID-19 government interventions. Nature Human Behavior 4:1303–1312. 14

[15] Kermack W.O., McKendrick A. G. (1927): A Contribution to the Mathematical Theory of Epidemics. Proceedings of the Royal Society of London. Series A, 115:700–721. 4

[16] Kermack W.O., McKendrick A. G. (1932): A Contribution to the Mathematical Theory of Epidemics II. The Problem of Endemicity. Proceedings of the Royal Society of London. Series A, 138:55–83. 4

[17] Kermack W.O., McKendrick A. G. (1933): A Contribution to the Mathematical Theory of Epidemics. III. Further Studies of the Problem of Endemicity. Proceedings of the Royal Society of London Series A, 141: 94–122. 4

[18] Khailaie, S., et al. (2021): Development of the reproduction number from coronavirus SARS-CoV-2 case data in Germany and implications for political measures. BMC Med, 19:32. 22

[19] Marshall, A. (2013[1890]): Principles of Economics. Palgrave Macmillan. ISBN 978-0-230-24929-5. 5

[20] May R.M., Anderson R.M. (1991): Infectious Diseases of Humans: Dynamics and Control. Oxford University Press. ISBN 0-19-854040-X. 2, 4

[21] Pullano, G., Di Domenico, L., Sabbatini, C.E. et al. (2021): Underdetection of Cases of COVID-19 in France threatens Epidemic Control. Nature, 590: 134–139. 22

[22] Ritchie, H., Mathieu, E. Rodés-Guirao, L., Appel, C., Giattino, C., Ortiz-Ospina, E., Hasell, J., Macdonald, B., Beltekian, D., Roser, M. (2020): Coronavirus Pandemic (COVID-19). Published online at OurWorldInData.org. Retrieved from: https://ourworldindata.org/coronavirus [Online Resource] 29

[23] Viceconte G., Petrosillo N. (2020): COVID-19 R0: Magic Number or Conundrum? Infectious Disease Reports, 12:8516. 4

[24] Wallinga J., Lipsitch M. (2007): How Generation Intervals Shape the Relationship between Growth Rates and Reproductive Numbers. Proceedings of the Royal Society of London Series B, 274:599–604. 7

[25] Weiss H. (2013): The SIR model and the Foundations of Public Health. MATerials MATemátics, Volume 2013, no. 3, 17 pp. ISSN: 1887-1097. 4

